# Negative predictive value Of Point-of-care ultrasound normal exam to exclude PneumoniA (NO-PNA)

**DOI:** 10.1101/2023.11.18.23298503

**Authors:** Larry Istrail, Shourjo Chakravorty, Maria Stepanova

**Affiliations:** Department of Medicine, Inova Fairfax Hospital, Falls Church, Virginia

**Keywords:** POCUS, point-of-care ultrasound, physical exam, Lung ultrasound, pneumonia

## Abstract

Pneumonia is one of the most common causes of hospital admission in the United States. Worldwide, the majority of antibiotics are prescribed for upper respiratory infections, much of which could be avoided with highly accurate methods of diagnosing or excluding pneumonia at the bedside. Despite the high prevalence of infectious pneumonia and decades of medical progress, accurately diagnosing or ruling out pneumonia without the use of computed tomography (CT) scanners and distinguishing between viral or bacterial etiologies remains challenging. We enrolled 61 patients in a prospective, blinded study to determine the sensitivity of lung ultrasound (LUS) using a handheld ultrasound probe in patients who had a computed tomography (CT) scan of their chest for any reason. The sensitivity of lung ultrasound to detect patients with evidence of pneumonia as compared to a CT scan was 94.4% (72.7 - 99.9) with a negative predictive value of 96.2% (80.4 - 99.9) when the scan was normal. Due to our broad inclusion criteria and the high prevalence of of non-infectious lung disease at the site of this study, the specificity was 61% (44.5 - 75.8) and positive predictive value was 51.5% (33.5 - 69.2). This simple lung ultrasound scan can be incorporated into the physical exam, has very high sensitivity to detect patients with pneumonia, and can be effectively used to rule out pneumonia if the lung exam is normal.

## Introduction

At its most basic level, pneumonia is an inflammatory process involving the alveolar space.^1^ While there are rare noninfectious causes, such as lipoid pneumonia or cryptogenic organizing pneumonia, most cases are caused by either bacterial or viral infections, which account for approximately 1.5 million hospitalizations each year in the United States.^2^ Despite the high prevalence of infectious pneumonia and decades of medical progress, accurately diagnosing or ruling out pneumonia without the use of computed tomography (CT) scanners and distinguishing between viral or bacterial etiologies remains challenging.

Pneumonia is often referred to as a clinical diagnosis,^3^ however physical exam findings in pneumonia are neither sensitive nor specific.^4,5^ This creates diagnostic uncertainty and, especially in low resource settings or outpatient clinics where immediate imaging is unavailable, incentivises antibiotic overprescription for respiratory tract infections.^6^ This is seen in both the United States and Europe.^7,8^ In the UK, 80–90% of all antibiotics are prescribed by general practitioners and most are prescribed for respiratory tract infections which are predominantly viral in nature.^8^ Overprescription of antibiotics leads to higher levels of microbial resistance and puts patients at risk for adverse reactions. These adverse reactions account for approximately 20% of drug related emergency department visits in the United States,^9^ and can range from allergic reactions to Clostridium difficile infections.^10^

When available, chest X-rays (CXR) are widely considered the radiographic standard for diagnosis, though their sensitivity is low when compared with CT. CXR can miss over 30% of infiltrates detected on CT, a poor sensitivity that was seen in multiple clinical settings.^11–14^ In a large retrospective study of 3,423 patients, CXR was only 43% sensitive for detecting pulmonary opacities seen with CT.^15^

Efforts to curb inappropriate antibiotic prescription trends generally rely on educational initiatives and clinician outreach,^16^ yet ultimately it is a problem of diagnostic uncertainty. Point-of-care testing, such as C-reactive protein (CRP) or procalcitonin levels, have been used to assist in the diagnosis of pneumonia as well. However, CRP is not sensitive or specific enough to eliminate uncertainty and only marginally improves the ability to diagnose pneumonia.^17^ When compared to CT confirmed lung consolidations in adult patients presenting with clinical signs of pneumonia, procalcitonin and CRP are not sensitive or specific enough to establish or rule out a pneumonia diagnosis, nor are they able to adequately differentiate bacterial from viral infections.^18^

Point-of-care ultrasound (POCUS) is another method for diagnosing pneumonia and other lung pathologies. In contrast to other point-of-care testing for non-specific biomarkers, lung ultrasound (LUS) offers the direct evaluation of the lung parenchyma for evidence of pneumonia. In a wide variety of populations, from neonates to the elderly, sensitivity and specificity for diagnosing pneumonia ranges from 85-95%. This was corroborated by multiple meta-analyses,^19–21^ which suggests POCUS could exclude pneumonia with a high degree of certainty. Abnormal lung ultrasound can also be suggestive of viral or bacterial etiologies, with larger lobar consolidations more specific for bacterial pneumonia, while distributed patches of subcentimeter consolidations and irregular pleura being more consistent with viral pneumonia.^22–25^

While this diagnostic accuracy is impressive, most studies were done using large, expensive cart-based hospital ultrasound machines in the intensive care unit or emergency department and their external validity in other clinical settings is less well understood. We sought to determine:

1) What is the sensitivity and negative predictive value of handheld lung ultrasound to detect patients with evidence of pneumonia as compared to CT scans in patients admitted to the hospital who had CT imaging for any reason?
2) What is the overall diagnostic accuracy and potential pitfalls in relying on ultrasound for diagnosing pneumonia?

## Methods

A prospective blinded study was completed to compare the ability of a standard lung ultrasound protocol using a handheld, smartphone-connected ultrasound probe (Butterfly IQ+, manufactured by Butterfly Network) to rule out pneumonia as compared to a gold standard CT scan of the chest.

Patients admitted to the hospital who underwent CT of their chest for any reason were recruited for this study if they had a BMI of 35 or under. Patients were excluded from the study if they were 1) intubated or on positive pressure ventilation 2) Had dressings or wounds preventing a full ultrasound exam of the thorax, or 3) Had chest tubes present. All patients provided written informed consent before enrollment in the study. The study was approved by the Institutional Review Board.

All ultrasound scans were performed by one physician (L.I.) certified in POCUS by the Society of Hospital Medicine and CHEST with 6 years of POCUS experience. In order to maintain blinding to the patient’s clinical information, type of CT scan, and reason for CT scan, a report of patients who had a chest CT scan of any kind in the 36 hours prior was made in the EPIC electronic medical record and downloaded into a spreadsheet. This spreadsheet only had the patient name, medical record number, BMI, bed location, and time of CT scan. L.I. was blinded to type of CT scan, admitting diagnosis, and all other medical information.

The accuracy of detecting pneumonia with POCUS, with CT-based pneumonia diagnosis used as a standard, was quantified via sensitivity, specificity, positive predictive value (PPV) and negative predictive value (NPV).

### Scanning protocol

The Butterfly IQ+ probe was initially placed in the “Lung” preset. Each lung was divided into six sections: two anterior, two lateral, and two posterior as described previously.^26^ The patient was placed in a supine position when evaluating the anterior and lateral lung sections and asked to sit up or roll to their side to assess the posterior sections.

The probe was placed in each section in a longitudinal plane and the probe was slid cranially and caudally, as well as laterally and medially in each section, looking for presence of A-lines, B-lines, lung sliding, pleural irregularities, pleural effusions, or consolidations. A clip was recorded in each section. For the lower lateral section and posterior inferior section, the liver and diaphragm were identified to confirm the base of the lung could be adequately visualized. When the lung was poorly visualized, the Butterfly IQ+ probe was switched to the “Abdominal” present to try to optimize the image.

A normal lung exam was deemed present in each section if there was presence of A-lines and lung sliding. An exam was deemed abnormal and consistent with a possible lung consolidation or pneumonia if any of the following were detected (Figure 1):

1. Localized, pathologic B lines defined as 3 or more B-lines in a given rib space
2. Jagged, irregular pleura with associated B -lines or subpleural consolidation
3. Confluent B lines in a given rib space
4. Subpleural (< 1cm) or large ( > 1cm in any dimension) consolidation

**Figure 1:**
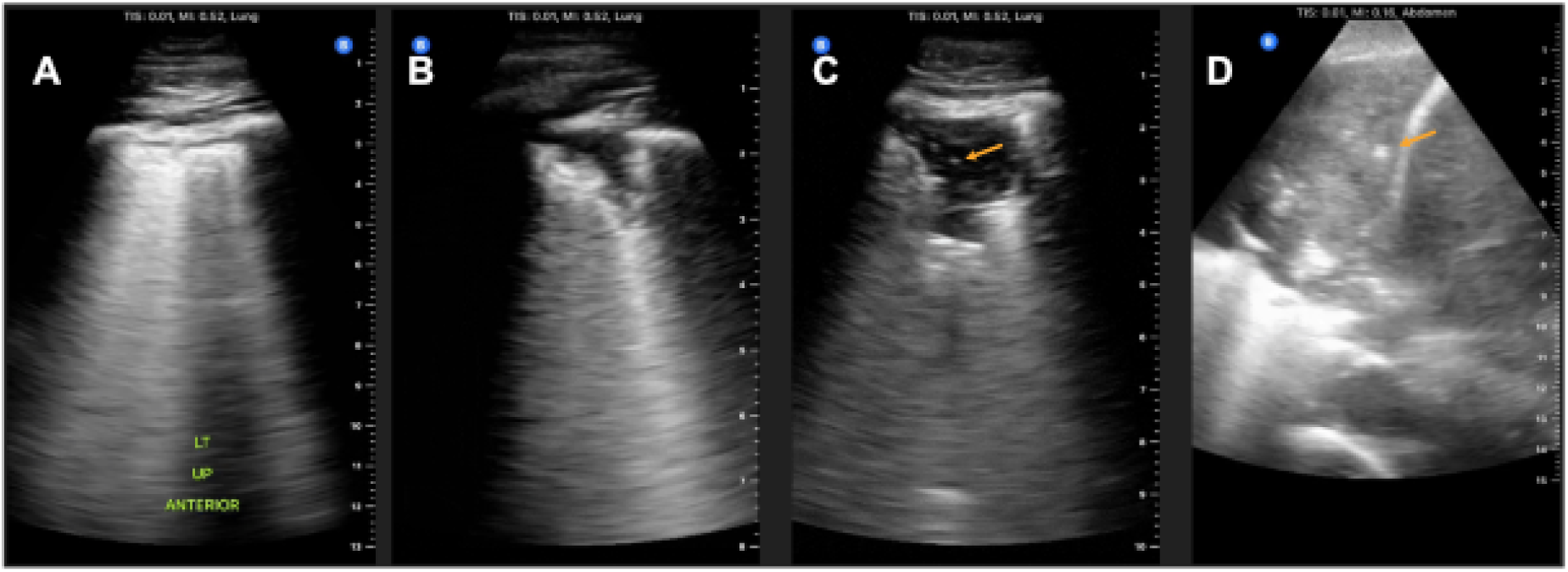
Spectrum of pneumonia. **A.** Pathologic B-lines with irregular pleura; **B.** Subpleural hypoechoic consolidation with irregular border < 1cm in size; **C.** Subpleural hypoechoic consolidation > 2cm in size with air bronchograms present; **D.** Dense consolidation with hepatization and air bronchograms; Air bronchograms labeled with orange arrow.

Findings were recorded on a paper worksheet and ultrasound clips were labeled and saved by L.I. The results were also recorded in a REDCap database. Another physician (S.C.) who was blinded to all ultrasound findings reviewed the CT scans and radiologist reads and recorded the results in a separate REDCap form. Once all patients were enrolled and all lung ultrasounds were completed and saved, the lung ultrasound results in each section were compared to the findings from the chest CT.

## Results

61 patients were consented and enrolled. Two patients were excluded due to inability to visualize the lung (one for bilateral breast implants and one for large bandage over a port that covered the anterior chest wall), and 59 were included in the analysis (39 male and 20 female). The average age was 63.9 (+/- 17.2) and average BMI of 23.2 (+/- 4.7).

The sensitivity of lung ultrasound to detect patients with evidence of pneumonia as compared to a CT scan was 94.4% (72.7 - 99.9) with a negative predictive value of 96.2% (80.4 - 99.9) when the scan was normal. There was only one false negative case in a patient with mild endobronchial patchy airspace opacities. Due to our broad inclusion criteria and the high prevalence of of non-infectious lung disease at the site of this study, the specificity was 61% (44.5 - 75.8) and positive predictive value was 51.5% (33.5 - 69.2).

## Discussion

In this study, we found that using a handheld ultrasound probe that attaches to a smart phone in patients with BMI under 35 is highly sensitive for detecting patients with evidence of pneumonia. Using a 12 zone protocol described previously,^26^ lung ultrasound was 94.4% sensitive for identifying patients with lung consolidations and therefore had excellent negative predictive value of 96% to rule out patients with evidence of pneumonia when the lung ultrasound was normal. In this study there was only one false negative case in which the patient had endobronchial mucous plugging with mild patchy airspace opacities that did not reach the pleura, and therefore was not detectable by ultrasound. This is consistent with the prior research which found that 98% of lung consolidations abut the pleura, and therefore it is possible but unlikely to miss a lung consolidation if all lung zones are scanned and are normal.^27^ This negative predictive value is superior to other commonly used point of care tests when their accuracy is compared compared to CT scans (**Table 2**).

**Table 2.**
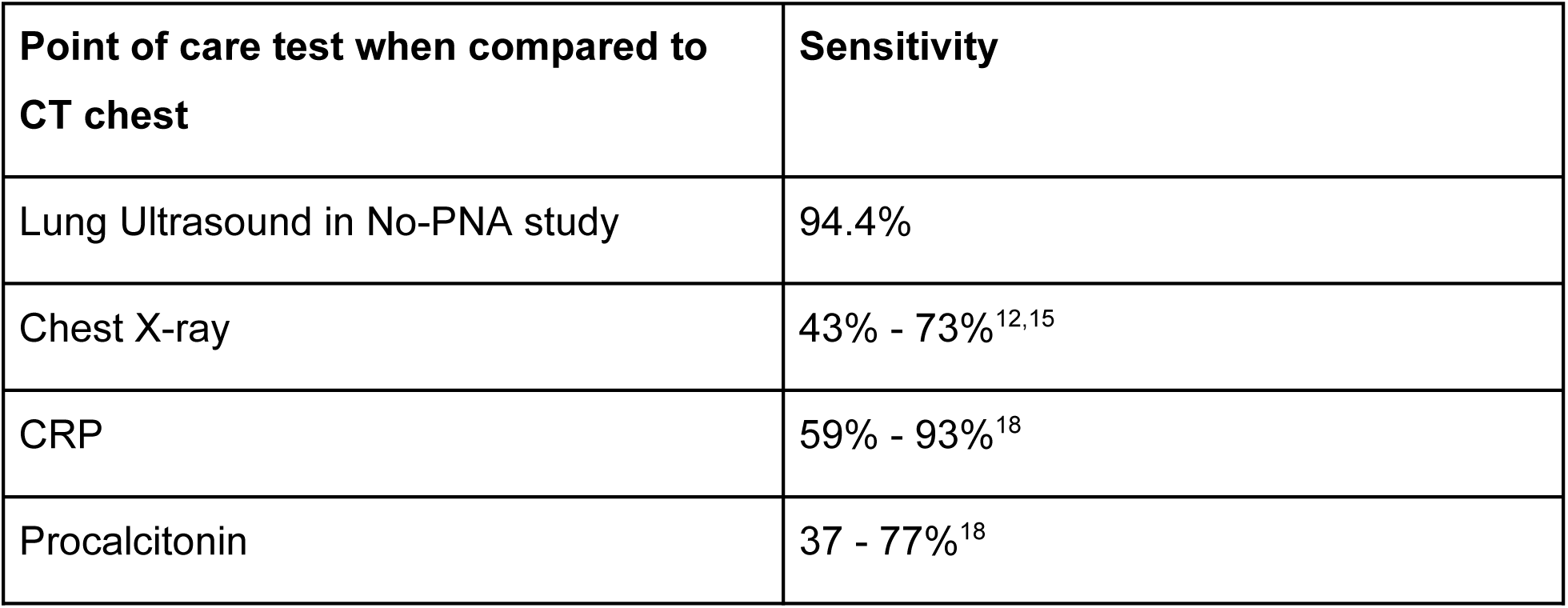
Sensitivity of Lung Ultrasound as compared to other diagnostic tests studied previously when compared with CT-scan proven lung consolidation.

| Point of care test when compared to CT chest | Sensitivity |
| --- | --- |
| Lung Ultrasound in No-PNA study | 94.4% |
| Chest X-ray | 43% - 73%<sup>12,15</sup> |
| CRP | 59% - 93%<sup>18</sup> |
| Procalcitonin | 37 - 77%<sup>18</sup> |

This study was designed to determine the sensitivity of lung ultrasound to detect any lung consolidation that could be consistent with viral or bacterial pneumonia when the scanner is completely blinded to all clinical information. We opted to enroll any patient who had a chest CT for any reason. This included chest CT angiography to evaluate for pulmonary embolus, high resolution CT chest, and CT chest with or without contrast. This was done to maintain blinding to any clinical information for the physician performing the lung ultrasound (LI), as well as to include all patients who had a CT of their chest regardless of the reason. Only enrolling patients with clinical evidence of pneumonia would likely improve the specificity but would lower the external validity when using lung ultrasound in patients with more vague respiratory symptoms. In addition to patients with pneumonia, by nature of our blinded enrollment process and patient population at our hospital system with a lung transplant service, our study subjects included patients with pulmonary fibrosis, cardiogenic pulmonary edema, pulmonary sarcoidosis, lung cancer, and pulmonary contusions after motor vehicle accidents, all of which cause lung parenchymal changes that can mimic the appearance of a lung consolidation related to pneumonia and reduce the speciticity of these findings.

The appearance of pneumonia varies if it is due to a viral, atypical bacterial, or typical bacterial infection, though there is significant overlap between all three. The use of lung ultrasound can also provide clues toward underlying cause of a particular pneumonia. Traditional bacterial pneumonia consolidations are more likely to be unilateral solitary consolidations greater than 2 cm in size with air bronchograms present.^25^ Dynamic air bronchograms are also sometimes present and are highly specific to pneumonia, though air bronchograms in lung consolidations consistent with pneumonia can be either static or dynamic.^28^

In contrast, viral pneumonia is characterized by patchy areas of ground glass opacities with pleural irregularities and possible subpleural consolidations, usually less than 1.5cm in size.^25,29^ This is also true of atypical pneumonias, such as those caused by *Mycoplasma pneumonia,*^30^ as well as non-infectious etiologies such as interstitial lung disease.^31^

When compared with CT as the gold standard, multiple meta-analyses have found lung ultrasound to be highly accurate in diagnosing pneumonia.^21,32^ This is especially true in children and younger populations without a large burden of fibrotic lung disease, where lung ultrasound accuracy approaches that of CT.^19^ In adults, Liu et al compared lung ultrasound with CT scans and had very high sensitivity of 94.6% and remarkably high specificity of 98.5%.^33^ They only included patients clinically suspected of CAP, and they defined lung ultrasound diagnosis of pneumonia as presence of any of the following: lung consolidation, focal interstitial pattern, ≥ 2 subpleural lesions, or ≥ 5 intercostal spaces with pleural-line abnormalities. They reported 100% specificity of two or more subpleural lesions for diagnosing CAP, a finding not consistent with our patient population. This is likely due to the fact that they did not enroll patients with pulmonary fibrosis, pulmonary sarcoidosis or lung cancer.

Amatya et al also had sensitivity over 90% but lower specificity of 61%.^34^ They enrolled patients with clinical suspicion of pneumonia and defined pneumonia as ‘the presence of unilateral B-lines or subpleural lung consolidations.’ False positives included tuberculosis, interstitial lung disease and bronchiectasis with fibrosis, which would explain their sensitivity and specificity similar to ours. Bourcier et al also had a lower specificity of 57% due to 9 false positive results from non-infectious pulmonary disease.^35^

In this study, in addition to false positives from pulmonary fibrosis or lung cancer, we encountered multiple cases where lung ultrasound detected subpleural consolidations consistent with pneumonia that corresponded with subtle findings on the CT not commented on in the radiology read. Many of these ultrasound findings were consistent with the final diagnosis despite a reportedly normal chest CT.

### Notable “false” positive cases

#### Case 1

Elderly female with history of rheumatoid arthritis and Sjogren’s syndrome on methotrexate presented with fever, cough and mild hypoxic respiratory failure. She had a temperature of 102.4, a white blood cell count of 11 x 10^9^/L, and Influenza and SARS-CoV2 were negative. Her chest CT read by radiology as having no acute consolidations. LUS revealed multiple patches of B lines with irregular pleura and sub-centimeter subpleural consolidations most consistent with acute viral infection (Figure 2). She tested positive for acute human metaneumovirus on a respiratory viral panel two days later.

**Figure 2.**
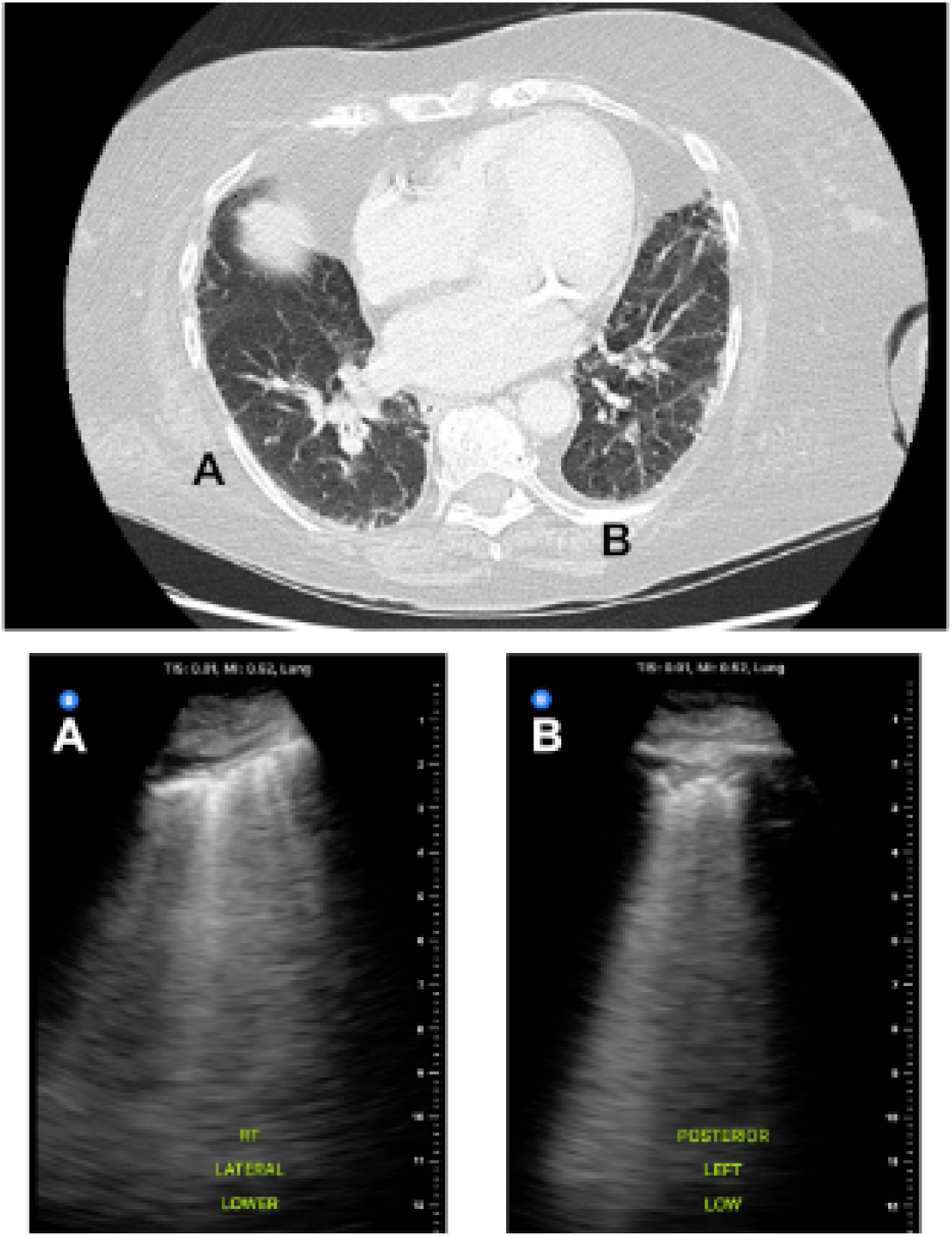
Pathologic B-lines with irregular pleura seen throughout posteriolateral lung fields. Two examples (**A, B**) which correspond to minimal changes on chest CT. Formal CT read by radiologist as no consolidations or acute pathology.

#### Case 2

Middle aged female with no medical history presented with fevers, malaise and cough. Her son tested positive for influenza A a few days prior to her admission. Her symptoms did not improve and she presented to the hospital. She was afebrile with white blood cell count of 10.96 x 10^9^/L. Respiratory viral panel, influenza and SARS-CoV2 were negative. CT chest was read as normal. However, ultrasound showed two areas of sub-centimeter subpleural consolidations with surrounding B-lines with irregular pleura consistent with a viral pneumonia that corresponded to subtle abnormalities seen on CT but not commented on (Figure 3).

**Figure 3:**
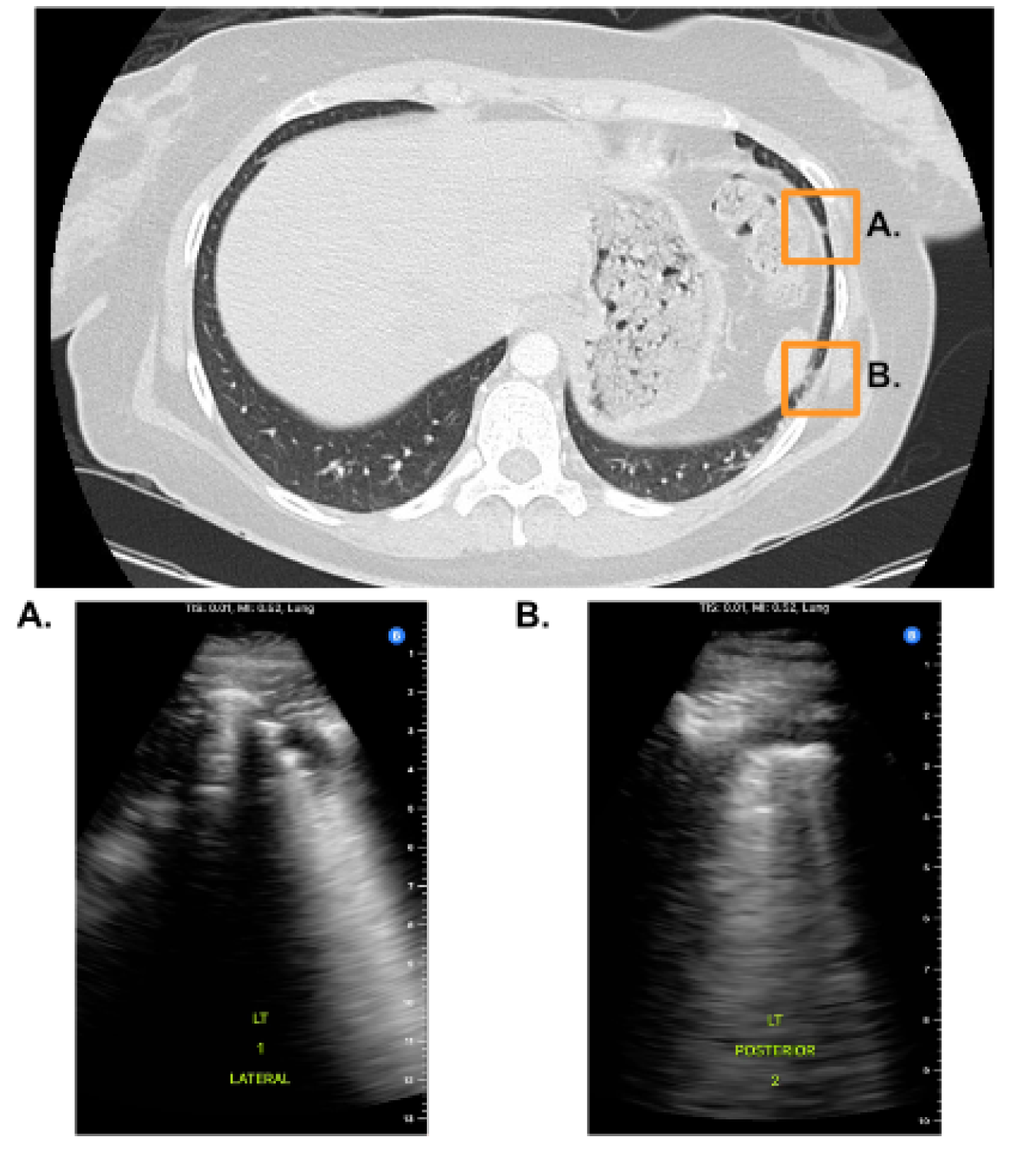
**A.** 1cm Subpleural consolidation with irregular pleura and focal, pathologic B-lines; **B.** Irregular pleura with associated pathologic B-lines. These findings together provide strong evidence of viral pneumonia in a healthy young female with no pulmonary disease despite normal CT scan read.

### Limitations

This study had multiple limitations. Despite all efforts to maintain blinding, once inside the patient’s room in some cases it was impossible to ignore what was hanging from an IV pole or whether or not the patient was receiving supplementary oxygen. With the handheld probe, resolution of the pleura was sometimes poor, especially in patients with thick subcutaneous tissue or who are obese. This limited the ability to determine how irregular or thickened the pleura actually was in some patients. The accuracy of LUS was also limited by the subjectivity that is inherent in interpreting chest CT scans. As shown in the highlighted cases, LUS sometimes appeared to correlate better with the clinical scenario than CT.

## Conclusion

In summary, LUS is a powerful tool that can be utilized to evaluate patients with respiratory complaints. Even with the lowest priced ultrasound probe available, the sensitivity of LUS was high enough to rule out pneumonia with nearly the certainty offered by a CT scan. This is especially useful in the outpatient and low resource settings, where it can have significant cost savings and eliminate or reduce the need for further bloodwork or imaging. LUS can also help differentiate viral vs bacterial pneumonia. These results may assist clinicians in making more informed decisions when evaluating patients with potential pneumonia and could reduce unnecessary antibiotic prescriptions.

## Data Availability

All data produced in the present study are available upon reasonable request to the authors

